# Planetary Health, Indigenous Knowledge and Healthcare: a Scoping Review

**DOI:** 10.1101/2022.11.28.22282853

**Authors:** Jihae Abou El Ela, Mercedes Mudgway, Niki Harré, James Hamill

**Affiliations:** Department of Paediatric Surgery, Starship Children’s Hospital; School of Psychology, the University of Auckland; Department of Paediatrics, Child and Youth Health, the University of Auckland, and the Department of Paediatric Surgery, Starship Children’s Hospital

**Author notes:** **Correspondence** James Hamill, Paediatric Surgery, Starship Children’s Hospital, Park Road, Auckland 1142, Aotearoa New Zealand.

## Abstract

**Background:** Planetary health is a movement to promote a healthy planet as the most important determinant of human health. Indigenous knowledge often encompasses a holistic view of nature and people in a way more akin to planetary health than to healthcare systems based on people alone. A change in healthcare’s worldview could help it become more environmentally sustainable. The aim of this review was to identify gaps in the literature and opportunities for further research at the nexus of Indigenous knowledge, environmental sustainability and healthcare.

**Methods:** We searched the databases Web of Science, Medline, and Google Scholar for peer-reviewed publications with terms pertaining to sustainability or planetary health, human healthcare, and indigenous or traditional knowledge. Papers were grouped by theme. Analysis was descriptive.

**Results:** The search process resulted in 10 eligible papers. Studies originated from 4 continents including one from Aotearoa New Zealand. Most (8/10) were published since 2020. Methodology included ethnography, discourse, imagery, descriptive, and quantitative analysis. One paper involved a particular healthcare system while all others considered general aspects of human health as related to the environment and Indigenous knowledge. Themes included 1) the importance of indigenous knowledge, 2) conceptual models that incorporate Indigenous approaches to health and the environment, and 3) the implementation of interventions. Within the limitations of the research available, it appears that Indigenous knowledge could make invaluable contributions to more environmentally responsible healthcare systems and can guide interventions to address planetary health problems.

**Conclusion:** Literature on Indigenous knowledge as related to planetary health and healthcare is limited and recent. There is scope for more research from many different Indigenous groups including Māori in Aotearoa, and scope for more collaborative research between healthcare systems and Indigenous peoples.

## Introduction

Healthcare plays in integral role in promoting the wellbeing of individuals and society but ironically, in doing so, it causes significant damage to ecology, contributes to climate change, and exacerbates social injustice.^1– 3^ The recognition that human health and the health of Earth are inextricably linked^4,5^ has led clinicians to create a movement for planetary health.^6,7^ Planetary health is defined as ‘the health of human civilisation and the state of the natural systems on which it depends’.^8^ Although the literature on planetary health has been substantial in recent time, much of it appears to come from a ‘modern’^9^ viewpoint in which the issue of climate change is viewed as a problem to be solved external to ourselves. By contrast, traditional viewpoints often regard the world and all living beings as interconnected^10^ as expressed by Al-Delaimy et al.:

> Earlier civilizations… consider the earth, nature, and its bounties a privilege to be cherished and respected because of the interdependence with the environment for their survival… They have ensured that they are part of the eco-system and are a positive force to live and let animals, plants and all other creations live.^11^

Interconnectedness is also explained in a statement made at the 23^rd^ World Conference on Health Promotion in 2019:

> Core features of Indigenous worldviews are the interactive relationship between spiritual and material realms, intergenerational and collective orientations, that Mother Earth is a living being – a ‘person’ with whom we have special relationships that are a foundation for identity, and the interconnectedness and interdependence between all that exists, which locates humanity as part of Mother Earth’s ecosystems alongside our relations in the natural world. ^12^

The burden of climate change does not fall equally on all. Indigenous populations are among one of the vulnerable groups that will be disproportionately burdened by the impacts of biodiversity loss, climate change, and rising sea levels.^13^ Reasons for this are multifactorial including a closer relationship with and dependence on the land, sea, and natural resources in combination with the social and economic inequities present in many Indigenous populations.^14^ Added to this is the growing body of evidence that Indigenous management and land tenure are key to safeguarding natural resources.^15^

Starship Children’s Hospital began addressing environmental issues through a sustainability program which included a research initiative to create a people-focused sustainability network based on previous work in education.^16^ Situated in Aotearoa New Zealand, our founding treaty, te Tiriti o Waitangi (the Treaty of Waitangi), mandates partnership between the two treaty partners. These are those of Māori ancestry and the British crown, with the latter usually interpreted today as non-Māori New Zealanders.^17^ Given the central part Indigenous peoples play in ecology and the sustainability problems in the health system, we set out to explore how Indigenous views and healthcare came together in the context of promoting environmental sustainability in order to inform and guide our hospital’s sustainability program.

Therefore, the purpose of this review was to scope the international literature on scientific research at the nexus of the natural environment, Indigenous knowledge, and healthcare and to identify opportunities for future research.

## Methods

The review protocol was registered on the Open Science Framework (osf.io/r4yh2). The present report conforms to the preferred reporting items for systematic reviews and meta-analyses extension for scoping reviews (PRISMA-ScR)^18^ and the enhancing transparency in reporting the synthesis of qualitative research (ENTREQ)^19^ as shown in Appendices 1 and 2.

Eligibility criteria included research papers that addressed Indigenous or traditional models or approaches to environmental sustainability and human health. Excluded were reviews, editorials, and opinion pieces. No date or language limits were applied.

Information sources for database searches were Web of Science, Medline, and Google Scholar. The search strategy was developed by two researchers (JA and JH) to reflect the three main concepts of the review: the natural environment, Indigenous knowledge, and healthcare. Combinations of key words and Medline subject headings were trialled in an iterative process to arrive at the final search strategies. The search strategy for Web of Science was as follows: TS=(sustainable-health OR planetary-health) AND (Indigenous OR Maori OR Aborig* OR Pacific-Island* OR Native-Americ* OR Indian OR Australia* OR New-Zealand* OR traditional-knowledge). The search strategy for Medline is available in Appendix 3. The last search was performed between 26 April and 11 May 2022.

Selection of sources of evidence was performed by two researchers (JA and JH). These reviewers drafted search strategies and performed the searches. The results were imported into Rayyan^20^, an application for performing systematic reviews. Duplicates were removed within Rayyan. The two reviewers screened titles and abstracts independently with blinding. Blinding was then removed and the reviewers resolved discrepancies by discussion and consensus. The two reviewers then screened the full texts independently with blinding, and then came together to resolve any discrepancies by discussion and consensus. In addition to the database search, we citation tracked, performed general searching of the internet, and pursued previous bibliographies compiled by one of the reviewers (JH) for relevant papers.

Data charting was by the same two reviewers who extracted data onto a spreadsheet after reading each paper multiple times. The data collection form was developed by two researchers (JA and JH). The data items included a description of outcomes and key findings related to the review, and the items presented in Table 1.

**Table 1.** Summary of studies included in the scoping review (in ascending order of date published).

| First author | Year | Country | Methodology | Quality score* |
| --- | --- | --- | --- | --- |
| Beaudin | 2010 | Canada | Ethnography | 90% |
| Kanene | 2016 | Zambia | Ethnography | 75% |
| DiPrete Brown | 2020 | Multiple | Workshops/expert | 60% |
| Jones | 2020 | Borneo | Quantitative and descriptive | 80% |
| Langmaid | 2020 | Australia | Interviews | 90% |
| Poland | 2020 | Canada | Story telling | 70% |
| Redvers | 2020 | Multiple | Story telling | 65% |
| Bradbey | 2021 | Australia & Aotearoa NZ | Descriptive | 55% |
| Smith | 2021 | Caribbean & Central America | Group sessions | 80% |
| Redvers | 2022 | Multiple | Groups, conference | 70% |

\* Quality score presented as percentages.

> Datasets were reviewed at a meeting of the reviewers and any discrepancies resolved by discussion and consensus.

Data items included article characteristics (author, country of origin, year of publication), the aims/purpose of the study (which was obtained either from the abstract or the introduction of the paper), the study population and sample size if the paper included it, the methodology if relevant, the study outcomes and details, and finally key findings that relate specifically to the scoping question.

Critical appraisal of included papers was by a customised checklist based on the Standards for Reporting Qualitative Research.^21^ Papers were scored as to whether they do or do not comply with each of the 20 items. Synthesis of results was descriptive within themes. Two reviewers read and re-read the papers, met and discussed the main focus and meaning of each paper, and by consensus developed theme headings for the purpose of descriptive analysis.

## Results

After duplicates were removed a total of 178 citations were identified from the database searches. Based on title and abstract screening, 141 were excluded leaving 37 to be retrieved and assessed for eligibility. Of these, 2 were excluded as the full text was not able to be retrieved, 22 were the wrong publication type (call-to-action, review article, commentaries without qualitative or quantitative data), and 5 were not assessing something within the research question. Through citation searching, we identified 30 articles, all of which were retrieved. Of these, 24 were excluded due to being the wrong publication type, and 4 were not assessing something within the research question. This left 10 papers eligible for this scoping review as shown in Fig. 1.

**Figure 1.**
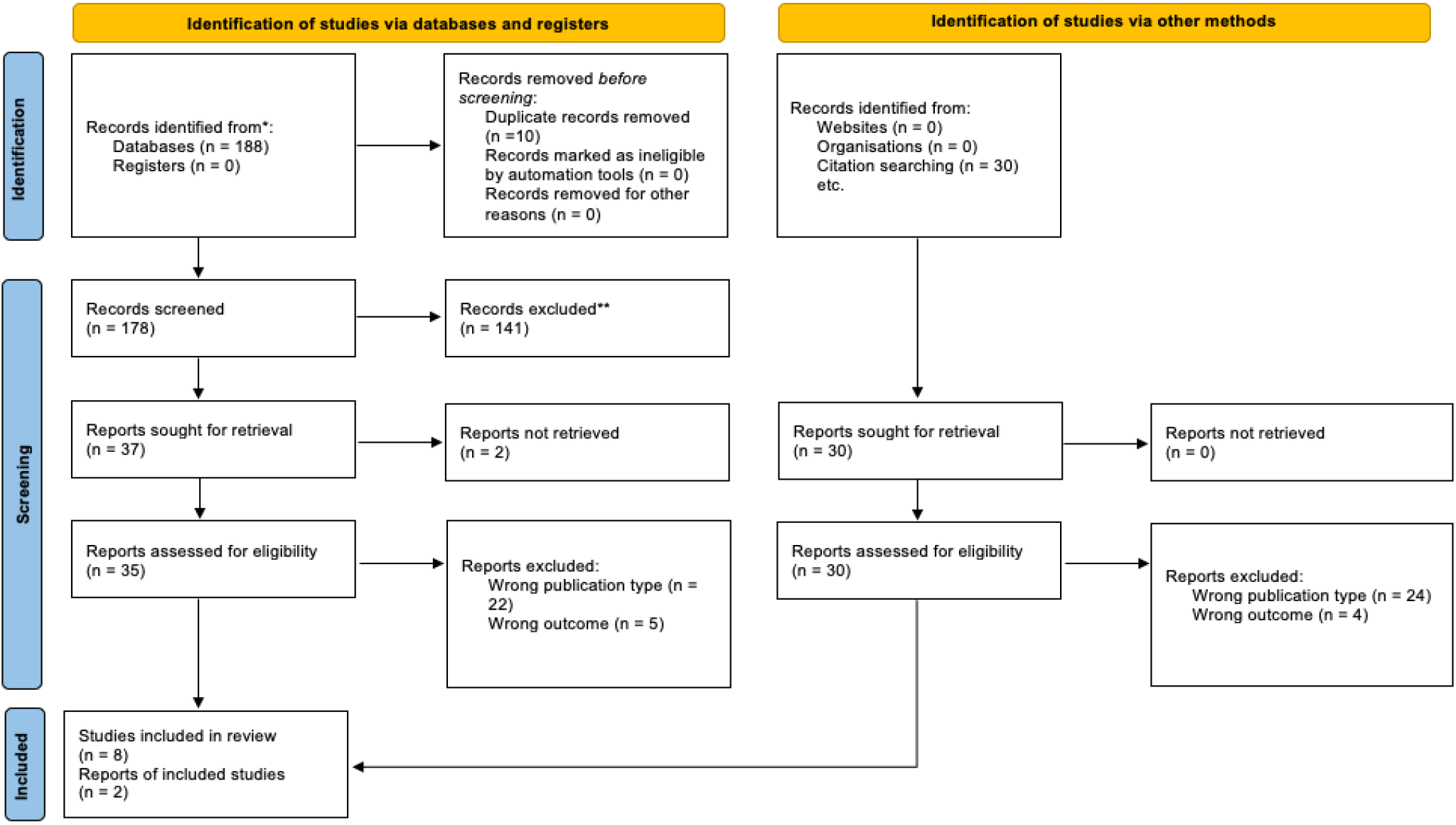
PRISMA flow diagram.^18^

Eight of the ten papers were published between 2020 and 2022. Geographically, research originated from Africa, Australia, Indonesia, Central and North America as shown in Table 1. The mean quality score of the studies was 14.5/20 (range 11 – 18, SD 2.4), as shown in Table 2. We classified papers into three main categories: 1) those that show the importance of Indigenous voices in planetary health; 2) papers that propose new models of indigenous worldviews, health and the environment; and 3) reports of interventions relating to Indigenous environmental health.

**Table 2.**
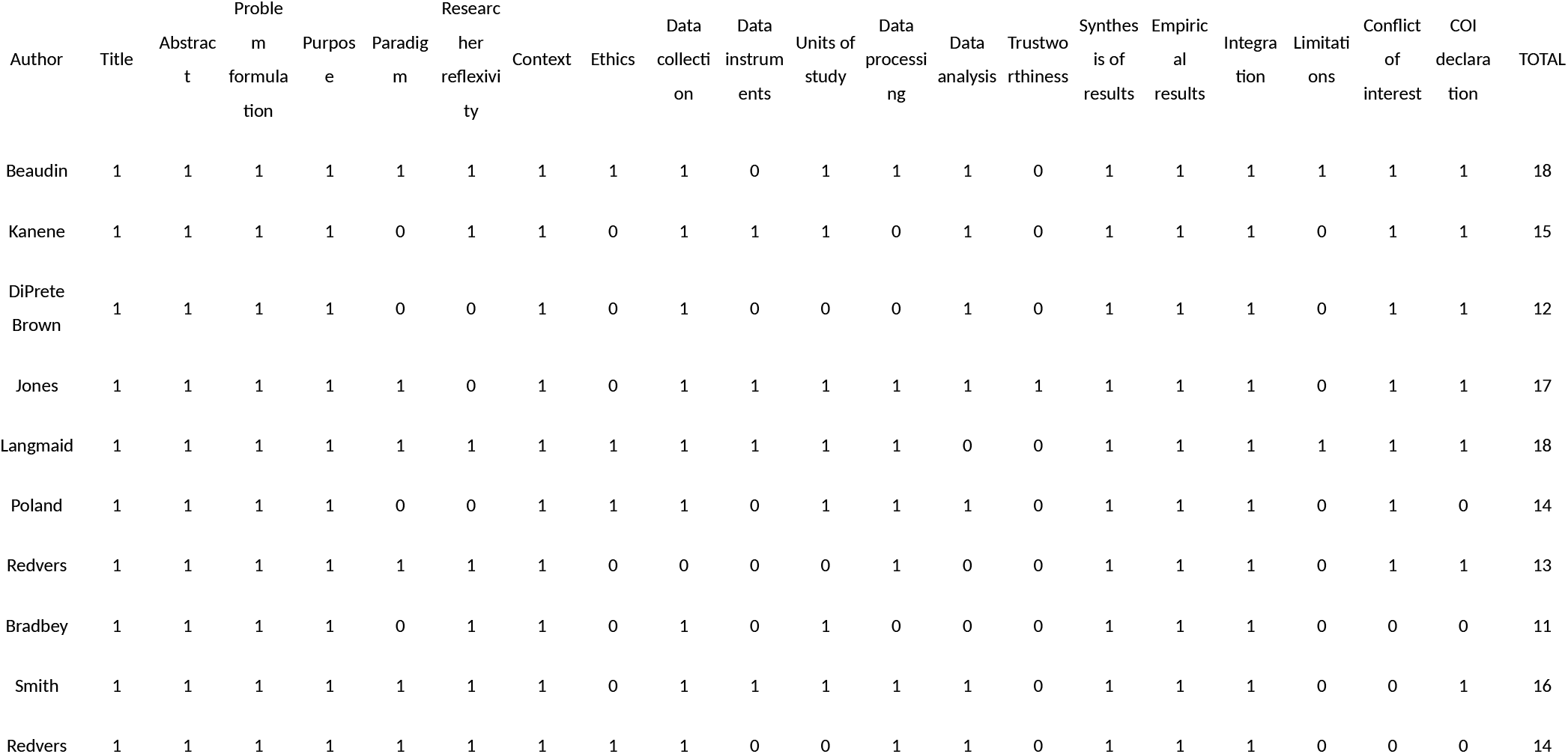
Quality assessment of the included studies. The total score was out of 20.

### Indigenous voices

Four studies focused on the lives and narratives of Indigenous peoples.^22–25^

An ethnographic study conducted by Beaudin in Canada focused on First Nations peoples’ perspective on health, healing and wellness.^22^ Beaudin found that a common theme throughout the community was that of connectedness between all things. Connectedness was experienced at different levels: within the community, between the community and the environment, and at the level of planet Earth. Connection promoted overall health and wellbeing at the physical, spiritual, mental and emotional level. Participants spoke of being stewards of the land and how changes to their environment − e.g. land contamination due to landfill sites near their community, or installation of power lines − influence individual and the community’s health.

> ‘… that’s our connection to Mother Earth, it’s our medicines. So our health has a lot to do with it, of who we are and what we believe in… when something happens to the land, it will affect us too’. ^22^

Kanene undertook an ethnographic study with the Tonga people of Southern Zambia.^23^ Trees and other plants were described as important sources of food, shade and windbreakers to the Tonga people. Certain trees are considered taboo and are therefore left untouched to the extent that not even fallen branches can be used for firewood. Extraction of nature-based medicines is done with consideration to environmental sustainability – only the required parts of the plant are taken because killing the plant is believed to equate to killing a human. Animals and birds are conserved and killing certain species is taboo. Water sources are considered habitats of god, thus no bathing, washing or dumping is permitted. Crop fields are not permitted along watercourses or dam catchment areas because eroded soils from the fields would result in the ‘death’ of the water source.

> The Tonga have cultivated and used biodiversity sustainably for centuries by supporting maintenance of healthy ecosystems.^23^

The studies by Beaudin and Kanene showed similar perspectives on the interconnectedness of nature and humans, on the effects of nature on human health and vice versa, and on environmental protection. Using a methodology based on imagery, Shelda-Jane Smith et al. show that the Maya communities of the Toledo District of Belize share similar concepts of the environment and wellbeing.^24^ The researchers asked participants to draw their vision of a healthy, sustainable future outside of colonial-liberal world views, and to describe their illustrations. Their overall findings are similar to the previously described studies. The authors summarised themes into two main concepts: 1) being children/people of the soil/earth, and 2) togetherness-community-dignity. Together they provide a framework for thinking about Indigenous Mayan health and wellbeing.

> What the Maya present is an understanding of health and wellbeing that is attained via strong, reciprocal relationships with land, forests, plants and people − factors which are frequently absent within Western accounts of health. ^24^

Poland et al. used talking circle methodology to learn the role and value of Indigenous knowledge traditions and practices in community engagement for envisioning sustainable futures.^25^ Participants came from the general public and student population in Toronto, Canada. Talking circles produced themes around relationships, spirit and sacredness, colonisation, and community. The authors found that traditional knowledge could have a role not only in environmental sustainability but in the way the research is conducted. They describe talking circles as a way to engage the population in conversation. The researchers relate Indigenous methodology to planetary health and to the ecological determinants of human health.

> Recognizing that problems cannot always be solved from the same level of thinking that created them, contemporary sustainability and social challenges are prodding us to look beyond conventional approaches to ‘greening’ business as usual… and to release the fiction that sustainability is a technical risk management problem, or even one of political will. In so doing, we create space to acknowledge the existence and relevance of alternative and hitherto marginalized worldviews, ways of knowing, and perspectives. ^25^

### Models

Four papers describe conceptual models that incorporate Indigenous approaches to the environment and human health.^26–29^

Langmain et al. introduce a revised version of a popular conceptual model adapted to fit with a planetary health mindset.^26^ The Mandala of Health,^30^ published in 1986 and used widely both in academia and practice, is described as favoured for its ‘ecological approach to wellbeing’. The original model displays a human at the centre and surrounded by the environment. This perhaps reflects a view of the environment as ‘separate, subserving and inferior to the human world…[with] topophylic, not biophilic connections to places’.^26^ Langmaid et al.’s modified Mandala of Health was guided by thematic analysis of interviews with ten ‘academics and practitioners working at the nexus of health promotion, environmental management and sustainability’.^26^ A key theme is ‘the cultural context of communities and gaining further understanding of Indigenous cultures are crucial elements of health promotion’.^26^ Indigenous concepts of interconnectedness are reflected in the core of the model featuring both human health and the health of the natural ecosystems to reflect the co-dependency of the two.

Redvers et al. used an Indigenous method of knowledge collecting to redefine the determinants of planetary health.^27^ The results show ten main determinants in three deeply interconnected levels. Similarly to Langmaid et al., this paper describes the ‘loss of awareness of the interconnectedness that exists within nature’ as one of the ‘pre-eminent causes of the planet’s destruction’,^27^ that is, the loss of an ecologically bound cultural identity. Redvers et al. also discuss the importance of epistemological pluralism, that is ‘an approach that recognizes that, in any given research context, there may be several valuable ways of knowing, and that accommodating this plurality can lead to more successful integrated study’.

Epistemological pluralism is further discussed in a second paper by Redvers et al. on molecular decolonisation in which they present the metaphor of a ‘two-eyed seeing lens’.^28^ Redvers et al. describe the impact that colonisation has at both a macro and micro level. The climate crisis is seen as a consequence of the appropriation of Indigenous land and the societal colonisation which causes disruption right down to the molecular level. The authors point out that ‘as a human species we all have the same molecular structure’.^28^ Disconnection has a negative impact on the planet; therefore, a process of decolonisation is required to promote interconnectedness and planetary health. Examples of ways to apply molecular decolonisation from a two-eyed seeing lens are fasting, reducing individual food consumption, and divesting from industrialised food systems, all of which can both reduce carbon footprint as well as improving individual human health. Other methods include restoring Indigenous cultural practices, language, and the art of storytelling as a method of sharing knowledge.

Models that support planetary health consider sustainability to be an integral foundational aspect rather than a ‘leg’ so to speak. DiPrete Brown et al. reflect this in their ‘three-legged stool’ model.^29^ Initially the three goals of sustainability − environmental protection, economic development and social development − are equated to a three-legged stool; however, this places humanity as separate from and outside the environment. With the environment being the floor, the authors suggest that the three legs should be 1) human rights and related legal frameworks, 2) gender analysis and gender mainstreaming practices, and 3) local and Indigenous history, knowledge and ways of knowing. These elements are ‘essential, complementary and congruent’ dimensions that can provide a balanced approach to any field interested in sustainable human development.

> Simply put, humanity can have neither an economy nor social wellbeing without the environment. Thus, the environment is not and cannot be a leg of the sustainable development stool. It is the floor upon which the stool, or any sustainable development model, must stand. It is the foundation of any economy and social wellbeing that humanity is fortunate enough to achieve.^29^

### Interventions

Two papers discuss the implementation of interventions aimed at promoting environmental and human health in Indigenous settings.^31,32^

Indonesia contains some of the most carbon-dense forests in the world. In Borneo in 2011, new logging concessions were temporarily banned in an effort to reduce total emissions from deforestation.^31^ This ‘top-down’ approach to the problem did not work as 61% of logging activity was illegal, and forests continued to be lost. Jones et al. initiated open-ended conversations amongst the rural Indigenous people to discover the drivers of illegal logging and develop interventions to improve access to healthcare. One intervention was to discount the cost of clinic visits in order to offset healthcare costs that were previously met through illegal logging. The research team also initiated education and alternative livelihood programs. Over 10 years, the program resulted in an estimated 70% reduction in deforestation and increased clinic usage and patient interaction. Early and continued collaboration with local communities is described as a key to the success of this intervention.

> Globally, about 35% of protected areas are traditionally owned, managed, used, or occupied by indigenous and local communities, yet the perspective and guidance of indigenous peoples and local communities is rarely considered in the design of conservation and climate mitigation programs.^31^

A second paper describes the Four Islands EcoHealth Network, a collaboration of initiatives across Australia, Tasmania, and Aotearoa New Zealand.^32^ The common factor amongst initiatives is the involvement of Indigenous peoples and communities, and the incorporation of traditional ecological knowledge. Bradby et al. suggest that ‘the restoration activities underway through our affiliated organizations… are having tangible benefits for human health’.^32^ While the paper presents little evidence for the link between the restoration activities described and health benefits, the authors acknowledge the need for further research on the health benefits of ‘expanded, holistic restorative activities at landscape, catchment, and regional scales’.^32^

## Discussion

This review found that the interaction between human health and the environment is a prominent feature of Indigenous knowledge. However, the number of included papers was surprisingly small leaving large gaps in the literature. Most papers were published since 2020 which suggests that interest in the nexus of health, the environment and Indigenous knowledge could be increasing.

Epistemologically, Indigenous peoples use oral traditions which represent collective knowledge acquired over thousands of years.^33^ In the Anthropocene, systems thinking is essential for navigating the complexities of earth and human behaviour. Holistic, systems thinking is typically characteristic of Indigenous worldviews.^34^ While ‘modern’ science traditionally tends to be more reductionist, there is an increasing emphasis on systems and networks.^35^ There is scope for more research where the investigators employ diverse epistemological positions.

The present review included papers from four continents but only one study involved Aotearoa New Zealand.^32^ Similar papers have been produced in Aotearoa but did not reach our inclusion criteria. Hall et al. described two projects for Indigenous partnership in urban ecosystem restoration in Aotearoa but did not specifically address health.^36^ Tu’itahi et al. described the importance of Indigenous worldviews as they relate to planetary health but their paper did not reach our criteria of research. There is scope for more research from Aotearoa and many other contexts worldwide.

Only one paper involved a particular healthcare system in primary health involving rural clinics.^31^ No paper addressed urban or hospital-based healthcare and Indigenous health and the environment. This is significant given the nefarious effect of the healthcare system on the environment. There is scope for more involvement of Indigenous peoples in reshaping the healthcare system into becoming ethical from a planetary health perspective.

Limitations of the present review include searching in a limited number of databases. We could have missed relevant papers, although the methodology was consistent with our aim of performing a scoping review.

Similarly, our search of grey literature was limited. The selection of papers that addressed human health involved judgement on the behalf of the reviewers. We sought papers that explicitly addressed some aspect of peoples’ health as well as environmental health. The quality measure was a non-validated customised version of a reporting guideline because standard measures of bias/quality are not designed for such a range of methodology as in the included papers.

In conclusion, literature at the nexus of health, the environment and Indigenous peoples is scarce. That which is available has highlighted the importance of traditional ecological knowledge and the value of Indigenous ways of being in approaching environmental and health concerns. There is scope for more research on all aspects of this topic and from a wider range of Indigenous peoples around the world.

## Data Availability

All data produced in the present work are contained in the manuscript

