## Appendix 2 for "Planetary Health, Indigenous Knowledge and Healthcare: a Scoping Review"

**Table 1 Enhancing transparency in reporting the synthesis of qualitative research: the ENTREQ statement**

| No | Item | Guide and description | Location in the manuscript |
|---|---|---|---|
| **1** | Aim | State the research question the synthesis addresses. | P 3 |
| **2** | Synthesis methodology | Identify the synthesis methodology or theoretical framework which underpins the synthesis, and describe the rationale for choice of methodology *(e.g. meta-ethnography, thematic synthesis, critical interpretive synthesis, grounded theory synthesis, realist synthesis, meta-aggregation, meta-study, framework synthesis).* | P 4 |
| **3** | Approach to searching | Indicate whether the search was pre-planned (*comprehensive search strategies to seek all available studies)* or iterative (*to seek all available concepts until they theoretical saturation is achieved).* | P 3 |
| **4** | Inclusion criteria | Specify the inclusion/exclusion criteria *(e.g. in terms of population, language, year limits, type of publication, study type).* | P 3 |
| **5** | Data sources | Describe the information sources used (e.g. *electronic databases (MEDLINE, EMBASE, CINAHL, psycINFO, Econlit), grey literature databases (digital thesis, policy reports), relevant organisational websites, experts, information specialists, generic web searches (Google Scholar) hand searching, reference lists)* and when the searches conducted; provide the rationale for using the data sources. | P 3 |
| **6** | Electronic Search strategy | Describe the literature search *(e.g. provide electronic search strategies with population terms, clinical or health topic terms, experiential or social phenomena related terms, filters for qualitative research, and search limits).* | P 3 & Appendix 3 |
| **7** | Study screening methods | Describe the process of study screening and sifting *(e.g. title, abstract and full text review, number of independent reviewers who screened studies).* | P 3-4 |
| **8** | Study characteristics | Present the characteristics of the included studies *(e.g. year of publication, country, population, number of participants, data collection, methodology, analysis, research questions).* | Table 1 |
| **9** | Study selection results | Identify the number of studies screened and provide reasons for study exclusion *(e,g, for comprehensive searching, provide numbers of studies screened and reasons for exclusion indicated in a figure/flowchart; for iterative searching describe reasons for study exclusion and inclusion based on modifications t the research question and/or contribution to theory development).* | P 4 & Fig. 1 |
| **10** | Rationale for appraisal | Describe the rationale and approach used to appraise the included studies or selected findings *(e.g. assessment of conduct (validity and robustness), assessment of reporting (transparency), assessment of content and utility of the findings).* | P 4 |
| **11** | Appraisal items | State the tools, frameworks and criteria used to appraise the studies or selected findings *(e.g. Existing tools: CASP, QARI, COREQ, Mays and Pope* [25]*; reviewer developed tools; describe the domains assessed: research team, study design, data analysis and interpretations, reporting).* | P 4 |
| **12** | Appraisal process | Indicate whether the appraisal was conducted independently by more than one reviewer and if consensus was required. | P 4 |
| **13** | Appraisal results | Present results of the quality assessment and indicate which articles, if any, were weighted/excluded based on the assessment and give the rationale. | Table 2 |
| **14** | Data extraction | Indicate which sections of the primary studies were analysed and how were the data extracted from the primary studies? *(e.g. all text under the headings "results /conclusions" were extracted electronically and entered into a computer software).* | P 4 |
| **15** | Software | State the computer software used, if any. | P 3 |
| **16** | Number of reviewers | Identify who was involved in coding and analysis. | P 4 |
| **17** | Coding | Describe the process for coding of data *(e.g. line by line coding to search for concepts).* | NA |
| **18** | Study comparison | Describe how were comparisons made within and across studies *(e.g. subsequent studies were coded into pre-existing concepts, and new concepts were created when deemed necessary).* | NA |
| **19** | Derivation of themes | Explain whether the process of deriving the themes or constructs was inductive or deductive. | NA |
| **20** | Quotations | Provide quotations from the primary studies to illustrate themes/constructs, and identify whether the quotations were participant quotations of the author's interpretation. | P 5-8 |
| **21** | Synthesis output | Present rich, compelling and useful results that go beyond a summary of the primary studies (e.g. *new interpretation, models of evidence, conceptual models, analytical framework, development of a new theory or construct).* | P 9-10 |

NA - formal coding and theme development was not applicable because this was a scoping review.
