## Appendix 3 for "Planetary Health, Indigenous Knowledge and Healthcare: a Scoping Review"

### Appendix 3. Search strategy for Medline

1. Indigenous Peoples/
2. Health Services, Indigenous/
3. Indigenous Canadians/
4. "Native Hawaiian or Other Pacific Islander"/
5. Indians, North American/
6. aborigin*.mp.
7. maori.mp.
8. historical.mp.
9. (Pre-industrial or Preindustrial).mp. [mp=title, abstract, original title, name of substance word, subject heading word, floating sub-heading word, keyword heading word, organism supplementary concept word, protocol supplementary concept word, rare disease supplementary concept word, unique identifier, synonyms]
10. oceanic ancestry group.mp.
11. indigenous.mp.
12. "Delivery of Health Care"/
13. (healthcare or health-care or health care).mp. [mp=title, abstract, original title, name of substance word, subject heading word, floating sub-heading word, keyword heading word, organism supplementary concept word, protocol supplementary concept word, rare disease supplementary concept word, unique identifier, synonyms]
14. "Environmental Restoration and Remediation"/
15. "Conservation of Natural Resources"/
16. Global Warming/ or Sustainable Development/ or environmental* sustainab*.mp. or Environmental Policy/
17. planetary health.mp.
18. or/1-11
19. or/12-13
20. or/14-17
21. and/18-20
